# Geographical validation of the Smart Triage Model by age group

**DOI:** 10.1101/2023.06.29.23292059

**Authors:** Cherri Zhang, Matthew O Wiens, Dustin Dunsmuir, Yashodani Pillay, Charly Huxford, David Kimutai, Emmanuel Tenywa, Mary Ouma, Joyce Kigo, Stephen Kamau, Mary Chege, Nathan Kenya-Mugisha, Savio Mwaka, Guy A Dumont, Niranjan Kissoon, Samuel Akech, J Mark Ansermino, the Pediatric Sepsis CoLab

## Abstract

Age is an important risk factor among critically ill children with neonates being the most vulnerable. Clinical prediction models need to account for age differences and must be externally validated and updated, if necessary, to enhance reliability, reproducibility, and generalizability. We externally validated the Smart Triage model using a combined prospective baseline cohort from three hospitals in Uganda and two in Kenya using admission, mortality, and readmission. We evaluated model discrimination using area under the receiver-operator curve (AUROC) and visualized calibration plots. In addition, we performed subsetting analysis based on age groups (< 30 days, ≤ 2 months, ≤ 6 months, and < 5 years). We revised the model for neonates (< 1 month) by re-estimating the intercept and coefficients and selected new thresholds to maximize sensitivity and specificity. 11595 participants under the age of five (under-5) were included in the analysis. The proportion with an outcome ranged from 8.9% in all children under-5 (including neonates) to 26% in the neonatal subset alone. The model achieved good discrimination for children under-5 with AUROC of 0.81 (95% CI: 0.79-0.82) but poor discrimination for neonates with AUROC of 0.62 (95% CI: 0.55-0.70). Sensitivity at the low-risk thresholds (CI) were 0.85 (0.83-0.87) and 0.68 (0.58-0.76) for children under-5 and neonates, respectively. Specificity at the high-risk thresholds were 0.93 (0.93-0.94) and 0.96 (0.94-0.98) for children under-5 and neonates, respectively. After model revision for neonates, we achieved an AUROC of 0.83 (0.79-0.87) with 13% and 41% as the low- and high-risk thresholds, respectively. The Smart Triage model showed good discrimination for children under-5. However, a revised model is recommended for neonates due to their uniqueness in disease susceptibly, host response, and underlying physiological reserve. External validation of the neonatal model and additional external validation of the under-5 model in different contexts is required.

**Author summary:** Clinical prediction model has become evermore popular in various medical fields as it can improve clinical decision-making by providing personalized risk estimate for patients. It is a statistical technique that incorporates patient-specific factors to personalize treatment and optimize health resources allocation. Clinical prediction models need to be validated in a different setting and population, and updated accordingly to ensure accuracy and relevance in clinical settings. We aim to evaluate one such model currently being implemented at the outpatient pediatric department at multiple hospitals in Uganda and Kenya. This model has been incorporated into a digital platform that is used to quickly identify critically ill children at triage. After validating the model against different age groups, we found the current model is not well suited for neonates and thus attempted to update the model. Our study provides new insight into clinical variables that impact neonatal outcome and we hope to improve neonatal morality for low-resource settings.

## Introduction

Significant progress has been made to decrease overall under the age of five (under-5) child mortality since the 1990s, though is due to improved outcomes among the non-neonatal population (1). Low- and middle-income countries (LMICs), particularly in Sub-Saharan Africa and South Asia continue to contribute disproportionately to childhood deaths globally (1). Amongst the five million children under-5 that died in 2020, 2.4 million were neonates with infectious diseases such as diarrhea, lower respiratory tract infections, meningitis, and malaria being major contributors (2). Sepsis is a dysfunctional inflammatory pathway leading to infection, death, and co-morbidities account for the majority of emergency and acute care visits in LMICs (3).

Effective triage facilitates early identification of critically ill children in and can improve outcomes through case prioritization, as most in-hospital deaths in resource-poor settings occur within 24 hours of admission (4, 5). The Emergency Triage Assessment and Treatment (ETAT) guidelines have been developed by the World Health Organization (WHO) for the assessment, triage, and initial management of acutely ill children in resource-poor facilities (6). The complexity of ETAT requires clinical knowledge, extensive memorization, and repetitive training, making its implementation a challenge in an environment where patient burden and new-staff turnover are high (7, 8, 9). An alternate solution is using electronic platforms with or without clinical prediction models which use data-driven algorithms to prioritize care (10, 11). One such example is the Smart Triage model, a 9-predictor pediatric triage model that can be embedded into a digital triage platform (12). Despite current progress, these models cannot be widely disseminated due to their lack of generalizability and external validation (13, 14, 15).

Prediction models are increasingly being used for individualized decision-making and to inform service delivery planning in health care (14). However, these models need to be externally validated before implementation in clinical settings (14, 16, 17). External validation can bridge the gap between the development and implementation of prediction models to ensure the model’s reproducibility and generalizability (14). Whether a model should be re-derived or updated during external validation depends on its performance in the validation cohort, the availability of research resources, and the characteristics of participants in which the model will be applied (15). By keeping the same predictors, updating a model can maintain predictive performance without losing prior information captured in the original model (16). Thus, there is a clear need to optimize existing prediction models for new settings (15).

Geographical validation and subsetting of data are two ways to evaluate prediction models. This study conducted geographical validation of the Smart Triage model by combining data from three hospital sites in Uganda and two sites in Kenya. Subsetting the data measures model accuracy to ensure the model is applicable to different subgroups in a dataset as certain subgroup(s) can lead to skewed or inaccurate predictions. As age is an important factor in risk prediction, we aim to remove older age groups from the data to measure model accuracy. The neonatal period is recognized as the period associated with the highest clinical risk (18), likely due to differences in inciting infections, disease susceptibility, host response, and underlying physiological reserve (19). To optimize the performance of the risk prediction across ages we hypothesize that patients under one month of age (neonates) have different risks and physiology and therefore require a different model. We update the model for this age group through a sequence of model-updating procedures (20).

## Method

Model external validation and updating followed to Transparent Reporting of a multivariable prediction model for Individual Prognosis or Diagnosis (TRIPOD) guidelines on developing, validating, or updating a multivariable clinical prediction model (21).

### Study population and design

The Smart Triage model was developed based on a prospective baseline cohort study conducted between April 2020 – March 2021 at the outpatient pediatric departments (OPD) of Jinja Regional Referral Hospital (Jinja), a public hospital funded by the Uganda Ministry of Health. It is the largest referral hospital in Eastern Uganda and serves patients residing in Jinja and eight surrounding districts. Its OPD, which functions similarly to an emergency department in high-income countries, evaluates between 20 and 100 patients per day and has an admission rate of approximately 20%.

The model was externally validated by combining baseline datasets from three additional sites in Uganda: Gulu Regional Referral Hospital (Gulu), Uganda Martyrs’ Ibanda Hospital (Ibanda), and St. Joseph’s Kitovu Hospital (Kitovu), and two sites in Kenya: Mbagathi County Hospital (Mbagathi) and Kiambu County Referral Hospital (Kiambu), separately for different age groups (< 30 days, ≤ 2 months, ≤ 6 months, and < 5 years). Ethics approval was obtained from the institutional review boards at the University of British Columbia in Canada (ID: H19-02398; H20-00484), Kenya Medical Research Institute (ID: KEMRI/SERU/CGMR-C/183/3958), the Makerere University School of Public Health in Uganda (ID: SPH-2021-41), and the Uganda National Council for Science and Technology (ID: HS1745ES).

Data used for validation were prospective baseline cohort studies conducted from March 2021 – April 2022 at Gulu, December 2021 – May 2022 at Ibanda, December 2021 – June 2022 at Kitovu, February – December 2021 at Mbagathi, and March 2021 – December 2022 at Kiambu. Study nurses were recruited and trained to conduct study-specific procedures. They recruited and consented participants in the triage waiting area using a quasi-random sampling method based on time cut-offs, and collected health data. The OPD at Gulu, Ibanda, and Kitovu see approximately 150,000, 19,000, and 19,000 patients annually with an admission rate of 18%, 33%, and 28%, respectively. The two hospitals in Kenya receive approximately 20,000 patients per year with an admission rate of 7% to 10%.

### Sampling and Eligibility

The full details of study procedures are presented in previous publications (10, 12). Briefly, children under 12, 15, or 19 years of age seeking assessment for an acute illness at the pediatric emergency department of all hospital sites between 8:00 am and 5:00 pm were enrolled using time cut off sampling procedures. In addition to parental/caregiver consent, assent was obtained for children above eight years of age at Jinja and Gulu and 13 years of age in Kenya; although this study only uses under-5 data. Children at Ibanda and Kitovu were not individually consented as the program was implemented as a quality improvement program with a waiver of individual consent. Children presenting for elective procedures, scheduled appointments, or treatment of chronic illnesses were not eligible for enrollment.

### Data collection and management

Data collection at all sites followed the same procedure that were used to develop the initial model at Jinja (12). Data were collected using password-secured Android tablets and a custom-built mobile application with an encrypted database. The Masimo iSpO2® Pulse Oximeter (City, Country) with micro USB connector was connected directly to the tablet to collect pulse oximetry and heart rate, and the Welch Allen SureTemp 692 (City, Country) thermometer was used to measure core temperature. Data was uploaded directly from the Android tablets to REDCap (Research Electronic Data Capture) (23) and sent to the central study server at the BC Children’s Hospital Research Institute and KEMRI Wellcome Trust Research Programme office for Uganda and Kenya, respectively. After each upload, data on the tablets was automatically deleted. Standard operating protocols for data collection and management are available on the Pediatric Sepsis CoLab Dataverse (24).

### Primary Outcome

The composite endpoint composed of one or more of the following: hospital admission of 24 hours or more determined from hospital records, readmission within 48 hours of enrollment, and mortality including in-hospital or post-discharge. Admission, readmission, and in-hospital or post-discharge mortality status were confirmed by follow-up phone calls to the caregivers 7 days post-study enrollment (for non-admitted patients) or post-discharge. As a secondary analysis, the proportion of children with a composite endpoint was compared using Fisher’s exact test between those over six months of age to under-5 and those six months and under.

### Smart Triage Model

The multiple logistic model was derived using bootstrap stepwise regression method and based on clinical validity with nine predictor variables included in the final equation. The model was integrated into a mobile application with a built-in pulse oximetry application that can be connected to a sensor, providing a smart algorithm that detects disease severity or level of risk in a child presenting to the hospital. The mobile application sends data to an interactive dashboard that provides clinical measurements and triage data to physicians in real-time allowing for rapid identification and assessment of critically ill children (12, 22).

The nine predictors included in the model were the square root of age, to attempt to linearize the nonlinear relationship between age and risk, heart rate, temperature, mid-upper arm circumference (MUAC), transformed oxygen saturation (using the concept of virtual shunt (25)), parental concern, difficulty breathing, oedema, and pallor (S1 Appendix).

### Statistical analysis

#### Sample Size

Sample size was pre-determined at each site prior to enrollment. In Uganda, it was computed based on the formula N = (nx10)/I where N is the sample size, n is the number of candidate predictor variables, and I is the estimated event rate in the population. The Smart Triage model developed at Jinja has nine predictors and an admission rate of 20%; thus, requiring a minimum sample of 450 participants. In other Ugandan sites, admission rate was used to calculate approximate sample size needed since predictors were already determined. In Kenya, a four-step procedure was implemented in the pmsampsize R package (26) to determine the minimum required sample to perform model validation. An input C-statistic of 0.8, an admission rate of 0.05, a Cox-Snell R-sq of 0.0697 based on 0.05 acceptable difference in apparent and adjusted R-squared, 0.05 margin of error in estimation of intercept, and an event per predictor parameter of 7 were assumed.

#### Model Validation, calibration, and update

The Smart Triage model equation was applied to a combined dataset comprising of five hospital sites, separated into four age groups: < 1 month, ≤ 2 months, ≤ 6 months, and < 5 years. These age groups were chosen based on the WHO’s age classification for special statistics of infant mortality (26). The older age groups were subsequently removed from the data starting with > 6 months, then > 2 months, followed by >= 1 month until only neonates remained to assess the model’s performance in the younger population. We also performed validation using exclusive age groups: >6m to < 5y, >2m to 6m, and 1m-2m, and neonates only. The sample size for each age category was still sufficient following the N = (nx10)/I rule. The model was assessed for its overall performance, discrimination, and calibration (14, 28, 29). The overall performance of the model was assessed using Brier Score, ranging from 0 to 1, with smaller values indicating a better model. Discrimination was assessed using area under the receiver operating curve (AUROC) and visualized with receiver operating characteristic (ROC) curves. AUROC close to 1 indicates good discrimination, while AUROC close to 0.5 indicates an inability to discriminate (28). Calibration was evaluated via calibration plots of predicted versus observed outcome rates with a 45-degree line representing perfect calibration (29). A calibration intercept of 0 and a slope of 1 is considered ideal. The model was updated for neonates only through a series of steps from recalibration to model revision, including the original Jinja cohort. The first step was recalibration-in-the-large to address the difference in baseline risks by re-estimating the model intercept. The next step was logistic recalibration to re-estimate the intercept and slope. Finally, a model revision was performed by re-estimating all regression coefficients using the same set of predictors. Age was not square rooted due to narrower range. Each step was visually examined with a calibration plot along with AUROC, Observed/Expected ratio, and calibration intercept and slope. An internal validation was performed using a 10-fold cross-validation procedure applied to the entire dataset due to the smaller sample size of neonates. A pooled estimate of AUROC, sensitivity, specificity, and predictive values was computed to quantify predictive accuracy.

#### Risk stratification

Following our previously reported model development process (12), two new risk thresholds were selected for the new models to divide participants into three triage categories (emergency, priority, and non-urgent). The low-risk threshold was selected at 90% sensitivity to limit misclassification of emergency and priority cases as non-urgent (avoiding false negatives), while the high-risk threshold was selected at 90% specificity to limit misclassification of non-urgent or priority cases as emergency cases (avoiding false positives). A risk classification table was used to examine the accuracy of the updated model in classifying patients.

Missing values were very few and were imputed using median and mode for continuous and categorical variables, respectively. Analyses were conducted in Stata version 15.0/MP (StataCorp, College Station, TX), R version 4.1.3 (R Foundation for Statistical Computing, Vienna, Austria), and RStudio version 2022.2.3 (RStudio, Boston, MA).

## Results

### Participants

A total of 13285 participants were enrolled in the study with 11595 (87%) under-5 included in the analysis and neonates accounted for 404 (3.5%) of the under-5. %). There was a higher prevalence of males in all the age groups, with a ratio of roughly 53% vs 46% (Table 1). Approximately 9% of the participants under-five were admitted to the hospital and that proportion increased as younger age groups were considered as the denominator, reaching 26% in neonates. There was a statistically significant difference (p-value < 0.0001) in the proportion of participants with a positive outcome between those over six months of age and those six months and under. For all age groups, more than 90% of those admitted had a minimum length of stay of 24 hours and less than 1% were sent home and readmitted within 48 hours. Pneumonia and neonatal sepsis were the most common reasons for admission in participants under-5. Malaria was more common in older children 140 (13.9%) and no neonates were admitted with a malaria diagnosis.

**Table 1.** Participant characteristics.

|  | < 5 years, n (%) | ≤ 6 months, n (%) | ≤ 2 months, n (%) | < 1 month, n (%) |
| --- | --- | --- | --- | --- |
| **Total participant<sup>1</sup>** | 11595 | 2421 (20.9) | 886 (7.6) | 404 (3.5) |
| **Sex** |  |  |  |  |
| Male | 6161 (53.1) | 1298 (53.6) | 494 (55.8) | 216 (53.5) |
| Female | 5434 (46.9) | 1123 (46.4) | 392 (44.2) | 188 (46.5) |
| **Outcomes** |  |  |  |  |
| Admitted<sup>2</sup> | 1004 (8.7) | 273 (11.3) | 154 (17.4) | 105 (26.0) |
| Length of stay ≥ 24 h<sup>3</sup> | 979 (97.5) | 260 (95.2) | 144 (93.5) | 96 (91.4) |
| Readmitted<sup>3</sup> | 20 (2.0) | 6 (2.2) | 3 (1.9) | 2 (1.9) |
| Within 48 h<sup>3</sup> | 3 (0.3) | 2 (0.7) | 1 (0.6) | 1 (1.0) |
| Mortality<sup>3</sup> | 27 (2.7) | 17 (6.2) | 9 (3.5) | 6 (5.7) |
| Not admitted<sup>2</sup> | 10591 (91.3) | 2148 (88.7) | 732 (82.6) | 299 (74.0) |
| Returned and readmitted<sup>3</sup> | 41 (0.4) | 7 (0.3) | 4 (0.5) | 2 (0.7) |
| Within 48 h<sup>3</sup> | 28 (0.3) | 1 (0.05) | 4 (0.5) | 2 (0.7) |
| Mortality<sup>3</sup> | 23 (0.2) | 7 (0.3) | 5 (0.7) | 4 (1.3) |
| Composite endpoint <sup>2</sup> | 1037 (8.9) | 278 (11.5) | 156 (17.6) | 105 (26.0) |
| **Admission diagnosis profile<sup>4</sup>** |  |  |  |  |
| Malaria | 140 (13.9) | 6 (2.2) | 2 (1.4) | 0 |
| Pneumonia | 425 (42.3) | 119 (43.6) | 39 (27.1) | 13 (12.4) |
| Bronchiolitis | 12 (1.2) | 7 (2.6) | 4 (2.8) | 1 (1.0) |
| URTI (cold, flu, etc) | 22 (2.2) | 1 (0.4) | 0 | 0 |
| Reactive Airway disease/asthma | 1 (0.1) | 0 | 0 | 0 |
| Gastroenteritis/diarrhea | 67 (6.7) | 4 (1.5) | 2 (1.4) | 0 |
| Meningitis/encephalitis or other CNS infection | 16 (1.6) | 2 (0.7) | 1 (0.7) | 1 (1.0) |
| Skin/soft tissue infection | 5 (0.5) | 2 (0.7) | 0 | 0 |
| Measles | 1 (0.1) | 0 | 0 | 0 |
| Sepsis | 67 (6.7) | 13 (4.8) | 9 (6.3) | 5 (4.8) |
| Neonatal sepsis | 42 (4.2) | 41 (15.0) | 38 (26.4) | 35 (33.3) |
| Anemia | 57 (5.7) | 12 (4.4) | 6 (4.2) | 1 (1.0) |
| Malnutrition | 22 (2.2) | 3 (1.1) | 2 (1.4) | 0 |
| Other (e.g., fever, convulsions, Jaundice, Neonatal Jaundice etc.) | 107 (10.7) | 63 (23.1) | 41 (28.5) | 49 (46.7) |

Note: age groups are inclusive, i.e. < 5 years includes the younger age groups. <sup>1</sup> percentage is based on total number included in the study, <sup>2</sup> percentage based on total number in the age group. <sup>3</sup> percentage based on the total number in the outcome category per age group, i.e. admitted and not admitted. <sup>4</sup> percentage based on total number of admissions per age group.

### Model performance and risk stratification

The overall performance of the Smart Triage model was best when all age groups were included with a Brier score of 0.08. As older age groups were removed, the Brier score increased to 0.18 in neonates. The model achieved good discrimination for all age groups, except neonates, with AUROC values ranging from 0.81 (95% CI: 0.79-0.82) for under-5 to 0.70 (95% CI: 0.65-0.76) for two months and under (Fig 1). The model achieved poor discrimination for neonates with an AUROC of 0.62 (95% CI: 0.55-0.70). The calibration slope also decreased in younger age groups with a value of 0.78 for under-5 to 0.42 for neonates (Fig 2). The assessment of exclusive age groups (>6m to < 5y, >2m to 6m, and 1m-2m) also resulted in good discrimination with AUROC of 0.82, 0.85, and 0.83, respectively, for all age groups except neonates (S1 Figure). Calibration plots showed a similar phenomenon (S2 Figure). When comparing model performance between inclusive and exclusive groups, the inclusion of younger participants decreased the model performance.

**Fig 1.**
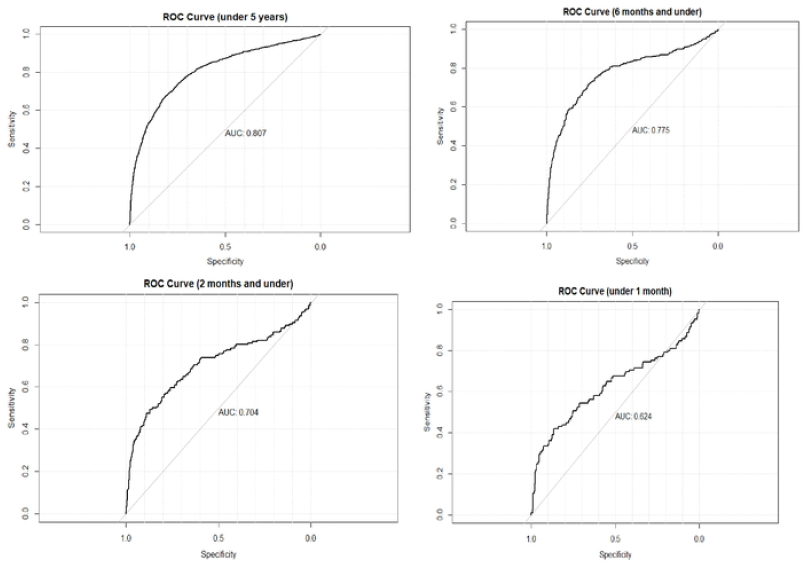
ROC curves by age groups.

**Fig 2.**
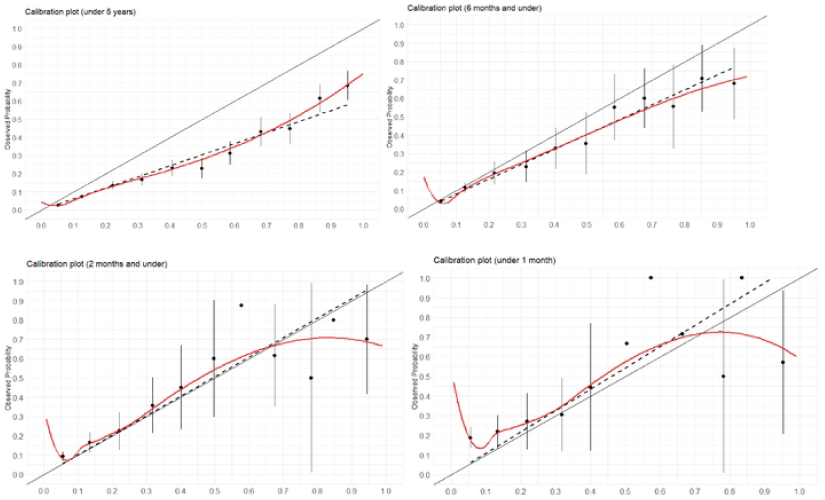
Calibration plots by age groups.

Table 2 shows the sensitivity and the specificity of the Smart Triage model by triage categories for each age group. The model achieved 85% (95% CI: 83%-87%) sensitivity at the low-risk threshold (non-urgent) and 93% (95% CI: 93%-94%) specificity at the high-risk threshold (emergency) for children under-5. Sensitivity decreased as older cohorts were taken out, while specificity remained relatively unchanged. For neonates, sensitivity was 68% (95% CI: 58%-76%) at the low-risk threshold, and specificity was 96% (95% CI: 94%-98%) at the high-risk threshold. The model identified around 10% of the participants as emergency. When examining mutually exclusive age groups, sensitivity remained high, ranging from 86% to 90% at the low-risk threshold, and specificity ranged from 93% to 98% (S1 Table).

**Table 2.**
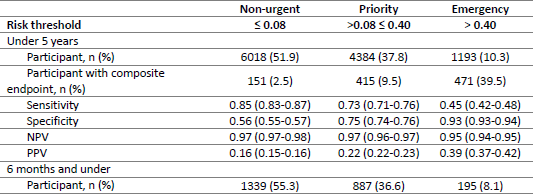
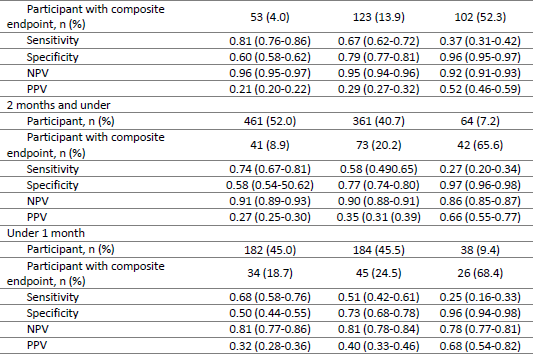
Summary of risk stratification into the three triage categories and model performance.

### Model update for neonates

As discrimination, calibration, and sensitivity dropped by 0.19, 0.36, and 17%, respectively, an updated model was developed for neonates. Fig 3 shows the sequence of the model update process. Each step resulted in an improvement in calibration (reaching the ideal value of 1 at step two) but limited improvement in discrimination until the final stage of model revision. The final model resulted in an AUROC of 0.83 (95% CI: 0.79-0.87) and calibration intercept and slope of 0 and 1, respectively (Fig 4). New intercept and coefficients for the predictor variables were derived. The equation from the updated model is:

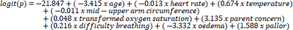

**Fig 3.**
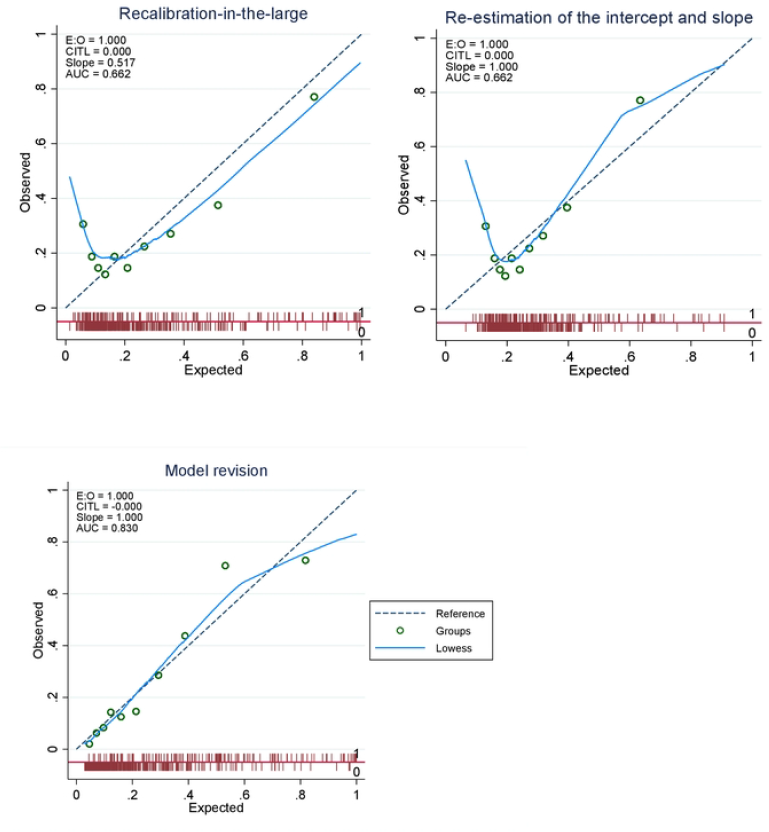
Model update for neonates only.

**Fig 4.**
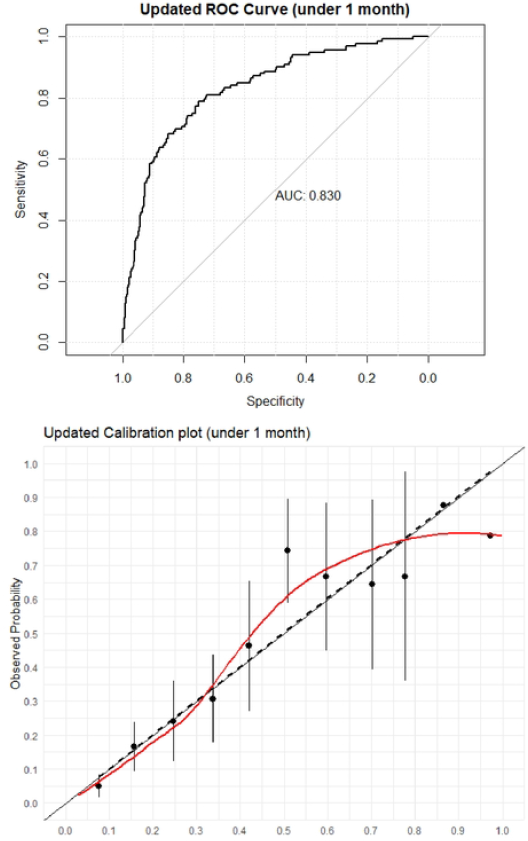
ROC curve and calibration plot of updated model for neonates.

Two new thresholds were chosen at 0.13 and 0.41 for the low and high risk, respectively, based on the desired sensitivity and specificity. Sensitivity at the low-risk threshold was 91% (95% CI: 86%-95%), and specificity at the high-risk threshold was 90% (95% CI: 87%-93%). The new model placed 23% of neonates into the emergency category, and 68% of those had an outcome.

**Table 4.** Summary of risk stratification in neonates by the updated model.

|  | Non-urgent | Priority | Emergency |
| --- | --- | --- | --- |
| Risk threshold | $\leq 0.13$ | $>0.13 \leq 0.41$ | $> 0.41$ |
| **Participant and Outcome Stratification** |  |  |  |
| Participant, n (%) | 178 (36.9) | 192 (39.8) | 113 (23.4) |
| Participant with composite endpoint, n (%) | 12 (6.7) | 43 (22.4) | 77 (68.1) |
| Admission status |  |  |  |
| Admitted, n (%) | 10 (5.6) | 46 (24.0) | 79 (69.9) |
| Not admitted, n (%) | 168 (94.4) | 146 (76.0) | 34 (30.1) |
| Length of stay |  |  |  |
| < 24h, n (%) | 0 | 5 (2.6) | 5 (4.4) |
| $\geq 24$h, n (%) | 10 (5.6) | 39 (20.3) | 74 (65.5) |
| Readmitted |  |  |  |
| Within 48 h, n (%) | 1 (0.6) | 1 (0.5) | 1 (0.9) |
| Beyond 48 h, n (%) | 1 (0.6) | 1 (0.5) | 1 (0.9) |
| Mortality, n (%) | 1 (0.6) | 6 (3.1) | 5 (4.4) |
| **Performance Assessment** |  |  |  |
| True positive to false positive ration | 120:185 | 106:98 | 78:35 |
| Sensitivity (95% CI) | 0.91 (0.86-0.95) | 0.80 (0.73-0.87) | 0.58 (0.49-0.66) |
| Specificity (95% CI) | 0.47 (0.42-0.52) | 0.72 (0.67-0.76) | 0.90 (0.87-0.93) |
| Negative predictive value (95% CI) | 0.93 (0.90-0.97) | 0.91 (0.88-0.94) | 0.85 (0.82-0.88) |
| Positive predictive value (95% CI) | 0.39 (0.37-0.42) | 0.52 (0.48-0.57) | 0.68 (0.61-0.76) |
| Negative likelihood ratio (95% CI) | 0.19 (0.10-0.33) | 0.28 (0.17-0.40) | 0.47 (0.37-0.59) |
| Positive likelihood ratio (95% CI) | 1.72 (1.48-1.98) | 2.86 (2.21-3.63) | 5.80 (3.77-9.43) |

## Discussion

This study externally validated the Smart Triage model in all children under five years of age using combined datasets from five hospital sites in Uganda and Kenya and evaluated performance across different age groups. Neonates had a higher proportion of composite endpoint at 26% compared to 8.9% in those under-5. The model achieved good discrimination with an AUROC of 0.81 in all children under-5. However, its discrimination decreased each time when older age cohorts were excluded, down to an AUROC of 0.62 for neonates. Calibration also deteriorated for those two months and under. Similarly, when comparing model performance using data that included neonates to data that did not, performance was better in the dataset that excluded neonates; therefore, the original Smart Triage model is not suitable for neonates. The model was updated for this age group by re-estimating the intercept and the coefficients of the selected predictors. Discrimination and calibration improved upon model revision with an AUROC of 0.83.

### External validation

External validation showed that the model maintained good discrimination in the new cohort at all sites. However, the model overpredicted the probability of having a composite endpoint due to a lower admission rate among the new cohorts at Gulu and the two sites in Kenya. Differences in the outcome incidence is the most likely cause of the miscalibration. When an algorithm is developed in a setting with high disease prevalence, it may systematically overestimate risk when used in settings with lower disease incidence (30). The difference in performance is due to the strength of the association between some predictors and the outcome being substantially different in the new population (31). This was evident when refitting the logistic regression in the new combined under-5 cohort (S2 Table). Heart rate and oedema were no longer statistically significant predictors because there were fewer percentage of children with oedema in the validation set despite the sample size being ten times larger than the derivation set (S3 Table). Additionally, the coefficient for square root of age changed from positive to negative in this refitting (S2 Table). The proportion of outcomes per categorical predictor were less in the external validation data set (S2 Table). Heterogenicity in predictor effects across the range of values of predictors can degrade overall performance. The discrimination and calibration decreased as older participants were excluded as would be clinically anticipated due to the differences in physiology and pathology in the younger age group. Another reason for the decrease in discriminative performance is that the range of age is smaller in neonates, and age is a predictor in our model that affects model performance (14, 32). Calibration in lower age groups is more heterogeneous (Fig 2), which may be magnified by smaller sample sizes, differences in case mix between the development and validation cohorts, and participants being clinically heterogeneous amongst each other.

### Model update for neonates

We updated the model for neonates as they differ in disease susceptibly, host response, and underlying physiological reserve. The first 28 days of life is the most vulnerable period and neonates are more likely to be admitted to hospital (18). Low birth weight, congenital malformations, unique infections, and gestational age are factors related to a worse outcome that may not significantly impact older children (33). Factors impacting neonatal outcomes may be different from those one month and older. This was evident in our data which indicated half of the predictors used in the original model to predict admission and mortality among children under-5 were no longer statistically significant in the updated model among neonates (S4 Table). Nonetheless, transformed oxygen saturation (OR 1.05, 95%CI 1.02-1.08) and temperature (OR 1.96, 95%CI 1.45-2.69) are still significant risk factors for a composite endpoint (S4 Table). The reduced performance of the model for neonates was anticipated. We followed a sequence of three updating methods that differed in extensiveness. Recalibration-in-the-large, adjusting the intercept (baseline risk), showed a modest improvement in calibration. Re-estimation of the intercept and slope continued to improve calibration; however, re-estimating all coefficients of the included predictors was necessary to improve discrimination as it is evident in the improvement of AUROC from 0.66 to 0.83. Previous studies have shown that when the model’s discrimination needs to be improved, revision methods are necessary (16, 32). Model updating with a new set of predictors may be the optimal approach; however, a trade-off between research resources and model improvement needs to be considered. Revision methods with small adjustments for predictors are preferred when a particular predictor in the original model has a different effect in the updating set (16). In our study, we opted to keep the original predictors and update the coefficients. A previous study has demonstrated that retaining the original selected variables where all coefficients are re-estimated using a large dataset improves both discrimination and calibration of the model (34).

### Clinical implication

External validation of clinical prediction models is critically important before implementation because models generally perform poorer during this external validation (14). It is imperative that prediction models are accurate in order to deliver precise case prioritization and appropriate individualized care recommendations based on illness severity to optimize patient outcomes. Thus far, a review that examined 84032 studies on prediction models concluded that only 5% had been externally validated (14). Furthermore, current neonatal predictive models used in low resources settings are developed mainly for estimating in-hospital mortality or limited to predefined populations such as infants born to mothers with severe preeclampsia or premature/low birth weight infants. We have demonstrated that the Smart Triage model, a parsimonious triage model, performs well during external validation in a different but related geographical context, making it reproducible. It is currently implemented at several sites in Uganda and Kenya (under review). We have also demonstrated that significantly improved performance can be achieved in a data subset by updating a model based on the same parsimonious variables. The high negative predictive performance of the model at the low-risk threshold demonstrates its ability to exclude low-risk patients. A positive predictive value of 68% (95% CI 61%-76%) was achieved at the high-risk threshold which, given the relatively low prevalence of composite endpoint, demonstrates the capability of the model to correctly classify emergency patients. The positive likelihood ratio indicates a 5.8-fold increase in the odds of needing hospital admission for a patient classified as emergency (Table 3). Furthermore, 91% of participants admitted for at least 24 h, and 92% of deaths were contained in either the priority or emergency category (Table 3). The parsimony of the Smart Triage model is easy to interpret and understand. This interpretability is particularly important where the results of a model may be used to make life-changing decisions. It requires less computation power and time to train, evaluate, and deploy compared to ETAT, making it practical for real-time applications where data may be limited and speed and efficiency are important.

### Strengths and Limitations

The key limitation of this study is that the validation sites were in adjacent geographical regions. The sites were similar in types of health facilities, but there were differences in disease prevalence and socioeconomic status between facilities that added to the heterogeneity of the cohorts. The optimal resemblance between the original and validation cohorts is a trade-off between reproducibility and generalizability. The next steps would be external validation on a different continent or in an urban tertiary care facility. Subsetting our data was a further limitation as subgroups in a dataset are underrepresented, leading to skewed or inaccurate predictions introducing a potential data bias. However, we used our clinical knowledge of age-specific disease processes to stratify our cohort into specific age categories, and with additional cohorts, we were able to stratify subsets by age and improve the model performance with model adjustment in the neonatal age group. A key strength of this study is the use of a large dataset from multiple sites for external validation to offer good statistical power. The use of routinely available clinical data resulted in low rates of missing data. The updated model was internally validated using the bootstrapping technique, which is the most widely recommended technique for internal validation as it allows derivation of the final model from the full derivation sample and does not waste precious information (35). In addition, the revised model used a portion of the original data, preventing overfitting (35).

## Conclusion

The Smart Triage model has been externally validated for similar clinical contexts in East Africa. An updated model for neonates is proposed, but will require additional external validation. The model is currently implemented in Uganda and Kenya to rapidly identify critically ill children at the first point of contact using routine clinical data and readily available vital signs. There is demonstrated evidence of improvement in the quality of care and patient outcomes as well as cost-effectiveness (36, 37). We believe the model is well adapted to use in resource-poor settings; however, further research is needed to continue to refine the model to increase its reproducibility and generalizability.

## Data Availability

The protocol, data dictionary, data collection software and code are currently unrestricted and available though the Pediatric Sepsis CoLab (https://doi.org/10.5683/SP3/MSTH98), subject to an application meeting the ethical and governance requirements of the CoLab (contact)

## Acknowledgement

We would like to thank the administration and staff of the Jinja Regional Referral Hospital, Gulu Regional Referral Hospital, Uganda Martyrs’ Ibanda Hospital, and St. Joseph’s Kitovu Hospital, Mbagathi County Hospital and Kiambu County Referral Hospital, and participants and caregiver for their time and dedication to Smart Triage. We would also like to thank Walimu and the Pediatric Sepsis CoLab.

## Supporting information

S1 Appendix. Smart Triage Model

S1 Figure. Calibration plots for exclusive groups

S2 Figure. ROC curves for exclusive groups

S3 Figure. 10-fold cross validated receiver operating characteristic curve for internal validation of the updated neonate model

S1 Table. Summary of risk stratification into the three triage categories and model performance

S2 Table. Comparison of logistic regression

S3 Table. Summary of categorical predictor variables stratified across derivation and validation set

## References

1. UNICEF. Under-five mortality 2023 [Available from: https://data.unicef.org/topic/child-survival/under-five-mortality/.

2. Organization WH. Child mortality (under 5 years) 2022 [Available from: https://www.who.int/news-room/fact-sheets/detail/levels-and-trends-in-child-under-5-mortality-in-2020.

3. Kwizera A, Kissoon N, Musa N, Urayeneza O, Mujyarugamba P, Patterson AJ, et al. A Machine Learning-Based Triage Tool for Children With Acute Infection in a Low Resource Setting. Pediatr Crit Care Med. 2019;20(12):e524-e30.

4. Kapoor R, Sandoval MA, Avendano L, Cruz AT, Soto MA, Camp EA, et al. Regional scale-up of an Emergency Triage Assessment and Treatment (ETAT) training programme from a referral hospital to primary care health centres in Guatemala. Emerg Med J. 2016;33(9):611-7.

5. Dekker-Boersema J, Hector J, Jefferys LF, Binamo C, Camilo D, Muganga G, et al. Triage conducted by lay-staff and emergency training reduces paediatric mortality in the emergency department of a rural hospital in Northern Mozambique. Afr J Emerg Med. 2019;9(4):172-6.

6. UPDATED GUIDELINE: Paediatric emergency triage, assessment and treatment care of critically ill children [press release]. 2016.

7. Molyneux E, Ahmad S, Robertson A. Improved triage and emergency care for children reduces inpatient mortality in a resource-constrained setting. Bull World Health Organ. 2006;84(4):314-9.

8. Hategeka C, Mwai L, Tuyisenge L. Implementing the Emergency Triage, Assessment and Treatment plus admission care (ETAT+) clinical practice guidelines to improve quality of hospital care in Rwandan district hospitals: healthcare workers’ perspectives on relevance and challenges. BMC Health Serv Res. 2017;17(1):256.

9. Mupara LU, Lubbe JC. Implementation of the Integrated Management of Childhood Illnesses strategy: challenges and recommendations in Botswana. Glob Health Action. 2016;9:29417.

10. Mpimbaza A, Sears D, Sserwanga A, Kigozi R, Rubahika D, Nadler A, et al. Admission Risk Score to Predict Inpatient Pediatric Mortality at Four Public Hospitals in Uganda. PLoS One. 2015;10(7):e0133950.

11. Mawji A, Akech S, Mwaniki P, Dunsmuir D, Bone J, Wiens MO, et al. Derivation and internal validation of a data-driven prediction model to guide frontline health workers in triaging children under-five in Nairobi, Kenya. Wellcome Open Res. 2019;4:121.

12. Mawji A, Li E, Dunsmuir D, Komugisha C, Novakowski SK, Wiens MO, et al. Smart triage: Development of a rapid pediatric triage algorithm for use in low-and-middle income countries. Front Pediatr. 2022;10:976870.

13. George EC, Walker AS, Kiguli S, Olupot-Olupot P, Opoka RO, Engoru C, et al. Predicting mortality in sick African children: the FEAST Paediatric Emergency Triage (PET) Score. BMC Med. 2015;13:174.

14. Ramspek CL, Jager KJ, Dekker FW, Zoccali C, van Diepen M. External validation of prognostic models: what, why, how, when and where? Clin Kidney J. 2021;14(1):49-58.

15. Binuya MAE, Engelhardt EG, Schats W, Schmidt MK, Steyerberg EW. Methodological guidance for the evaluation and updating of clinical prediction models: a systematic review. BMC Med Res Methodol. 2022;22(1):316.

16. Janssen KJ, Moons KG, Kalkman CJ, Grobbee DE, Vergouwe Y. Updating methods improved the performance of a clinical prediction model in new patients. J Clin Epidemiol. 2008;61(1):76-86.

17. Ewout W S. Clinical Prediction Models: A Practical Approach to Development, Validation, and Updating. Second ed. Gail M, editor. Switzerland: Springer; 2019.

18. UNICEF. Levels and Trends in Child Mortality. 2023.

19. Nemetchek BR, Liang LD, Kissoon N, Ansermino JM, Kabakyenga J, Lavoie PM, et al. Predictor variables for post-discharge mortality modelling in infants: a protocol development project. Afr Health Sci. 2018;18(4):1214-25.

20. Vergouwe Y, Nieboer D, Oostenbrink R, Debray TPA, Murray GD, Kattan MW, et al. A closed testing procedure to select an appropriate method for updating prediction models. Stat Med. 2017;36(28):4529-39.

21. Collins GS, Reitsma JB, Altman DG, Moons KG. Transparent reporting of a multivariable prediction model for individual prognosis or diagnosis (TRIPOD): the TRIPOD Statement. BMC Med. 2015;13:1.

22. Mawji A, Li E, Komugisha C, Akech S, Dunsmuir D, Wiens MO, et al. Smart triage: triage and management of sepsis in children using the point-of-care Pediatric Rapid Sepsis Trigger (PRST) tool. BMC Health Serv Res. 2020;20(1):493.

23. Harris PA, Taylor R, Thielke R, Payne J, Gonzalez N, Conde JG. Research electronic data capture (REDCap)--a metadata-driven methodology and workflow process for providing translational research informatics support. J Biomed Inform. 2009;42(2):377-81.

24. Mawji A. Smart Triage Jinja Standard Operating Protocols, V1 [dataset] Scholars Portal Dataverse2021 [Available from: https://borealisdata.ca/dataset.xhtml?persistentId=doi:%2010.5683/SP2/WLU0DJ.

25. Tushaus L, Moreo M, Zhang J, Hartinger SM, Mausezahl D, Karlen W. Physiologically driven, altitude-adaptive model for the interpretation of pediatric oxygen saturation at altitudes above 2,000 m a.s.l. J Appl Physiol (1985). 2019;127(3):847-57.

26. Ensor J, Martin EC, Riley RD. Package ‘pmsampsize’. 2022.

27. Nations U. Provisional guidelines on standard international age classifications. New York: United Nations, Department of International Economic and Social Affairs, Statistical Office; 1982. Contract No.: M.

28. Steyerberg EW, Vickers AJ, Cook NR, Gerds T, Gonen M, Obuchowski N, et al. Assessing the performance of prediction models: a framework for traditional and novel measures. Epidemiology. 2010;21(1):128-38.

29. Alba AC, Agoritsas T, Walsh M, Hanna S, Iorio A, Devereaux PJ, et al. Discrimination and Calibration of Clinical Prediction Models: Users’ Guides to the Medical Literature. JAMA. 2017;318(14):1377-84.

30. Steyerberg EW, Roobol MJ, Kattan MW, van der Kwast TH, de Koning HJ, Schroder FH. Prediction of indolent prostate cancer: validation and updating of a prognostic nomogram. J Urol. 2007;177(1):107-12; discussion 12.

31. Su TL, Jaki T, Hickey GL, Buchan I, Sperrin M. A review of statistical updating methods for clinical prediction models. Stat Methods Med Res. 2018;27(1):185-97.

32. Toll DB, Janssen KJ, Vergouwe Y, Moons KG. Validation, updating and impact of clinical prediction rules: a review. J Clin Epidemiol. 2008;61(11):1085-94.

33. Medvedev MM, Brotherton H, Gai A, Tann C, Gale C, Waiswa P, et al. Development and validation of a simplified score to predict neonatal mortality risk among neonates weighing 2000 g or less (NMR-2000): an analysis using data from the UK and The Gambia. Lancet Child Adolesc Health. 2020;4(4):299-311.

34. Cooray SD, Boyle JA, Soldatos G, Allotey J, Wang H, Fernandez-Felix BM, et al. Development, validation and clinical utility of a risk prediction model for adverse pregnancy outcomes in women with gestational diabetes: The PeRSonal GDM model. EClinicalMedicine. 2022;52:101637.

35. Massaut J, Valles P, Ghismonde A, Jacques CJ, Louis LP, Zakir A, et al. The modified south African triage scale system for mortality prediction in resource-constrained emergency surgical centers: a retrospective cohort study. BMC Health Serv Res. 2017;17(1):594.

36. Novakowski SK, Kabajaasi O, Kinshella MW, Pillay Y, Johnson T, Dunsmuir D, et al. Health worker perspectives of Smart Triage, a digital triaging platform for quality improvement at a referral hospital in Uganda: a qualitative analysis. BMC Pediatr. 2022;22(1):593.

37. Li ECK, Grays S, Tagoola A, Komugisha C, Nabweteme AM, Ansermino JM, et al. Cost-effectiveness analysis protocol of the Smart Triage program: A point-of-care digital triage platform for pediatric sepsis in Eastern Uganda. PLoS One. 2021;16(11):e0260044.

